# Using Natural Language Processing as a Scalable Mental Status Evaluation Technique

**DOI:** 10.1101/2023.12.15.23300047

**Authors:** Margot Wagner, Jasleen Jagayat, Anchan Kumar, Amir Shirazi, Nazanin Alavi, Mohsen Omrani

## Abstract

Mental health is in a state of crisis with demand for mental health services significantly surpassing available care. As such, building scalable and objective measurement tools for mental health evaluation is of primary concern. Given the usage of spoken language in diagnostics and treatment, it stands out as potential methodology. Here a model is built for mental health status evaluation using natural language processing. Specifically, a RoBERTa-based model is fine-tuned on text from psychotherapy sessions to predict mental health status with prediction accuracy on par with clinical evaluations at 74%.

## 1 Introduction

There is currently a mental health crisis as the demand for mental health services vastly outstrips the availability of quality care. In the United States and Canada, approximately 60 million people grapple with mental health concerns, yet regrettably, over two-thirds of those in need are unable to receive care despite a staggering $250 billion in healthcare expenditures [3, 2]. The prevailing model of care delivery relies on one-on-one interactions between clinicians and patients, which is labor-intensive and thus lacks scalability. The paucity of mental healthcare clinicians hampers the efficacy of this traditional format. The World Health Organization estimates around half the world’s population lives in countries where there is one psychiatrist for 200,000 or more citizens [1]. Furthermore, the considerable degree of clinician involvement further inflates the cost of conventional treatment. The COVID-19 pandemic has exacerbated this mismatch in care by increasing demand for services, fostering mental health awareness and diminishing the stigma associated with seeking care. This has created a persistent disparity between the supply and demand for mental healthcare, enduring even beyond the pandemic’s cessation. Consequently, it is imperative to prioritize the development of innovative solutions that streamline and automate the delivery of mental healthcare in order to address this public health concern.

The field of mental health care has lagged behind broader innovation in the healthcare industry. This discrepancy primarily arises from the dearth of robust and precise quantitative measurements, biomarkers, and evidence-based practices in this domain compared to other medical specialties.

Thus, it becomes essential for the field to center its efforts on the cultivation of novel data-driven measurement tools which can effectively evaluate the mental status of patients. Thomas Insel, former head of the National Institutes of Mental Health, argues that a new diagnostic system based on emerging research that incorporates multiple layers of information is a pressing need in psychiatry [17, 15]. By doing so, the aim is to construct comprehensive and objective protocols that are capable of adeptly handling mental health issues, thereby bridging the existing care gap and ensuring optimal treatment for individuals in need.

Establishing objective measurement tools for mental health evaluation is challenging due to their inherent complexity. Unlike other chronic diseases which typically utilize one or few biophysiological target variables, such as blood glucose in diabetes or blood pressure in hypertension, mental health disorders lack definitive biomarkers. As a result, there is a reliance on indirect measurements, including changes in sleep pattern or activity level [28] or changes in body posture [12] and speech tone [16]. While these biophysical symptom proxies hold significance in a clinician’s decision-making process, the patient’s thought content remains the most influential factor in diagnosis and treatment determination. The assessment of thought form and content, mood status, stressors and anxiety level predominantly relies on the patient’s verbal expression. Consequently, it becomes natural that the pursuit of a readily scalable technology for appraising a patient’s mental status would incorporate the patient’s linguistic utilization as a reliable information source. However, patients’ speech is unstructured data, which makes the process of extracting clinically relevant data a challenging endeavor. Even in clinical settings, therapists and clinicians must attentively and carefully listen to their patients to extract well-defined structured data suitable for diagnosis, intervention and monitoring purposes.

Deep learning methods have gained prominence in the field of mental illness detection, specifically in the task of natural language processing (NLP) [27, 10, 7, 4, 14]. Unlike traditional statistical and machine learning methods, deep learning methods do not heavily rely on feature engineering and can process longer, more complex sentences in a context-dependent manner. They also exhibit enhanced capabilities in learning languages structures, allowing for effective transfer learning with limited data. Transformers, newer than convolutional and recurrent neural networks, show promise in handling sequential and textual data, making them suitable for mental health applications[9]. In this work, the focus is on fine-tuning a pretrained transformer model to detect symptomatic sentences related to depression and anxiety in a client’s narrative. The paper acknowledges the state-of-the-art position of machine learning in NLP and includes a comparison with commonly used models in the results section.

## 2 Methods

### 2.1 Datasets

#### 2.1.1 Training Dataset

The non-clinical training data for this study was collected from online mental health forums where individuals share their personal experiences and challenges related to mental health. To prepare the data, the stories were segmented into sentences, and each sentence was carefully examined and labeled by an expert clinician (Expert A) as either neutral or exhibiting signs of anxiety and/or depression. In addition, sentences that relied on the previous or following sentence for context were flagged as dependent examples. An illustration of this dependency can be seen in the following pair of sentences: “Would I say my life is perfect and I am happy every day? No.”. In isolation, each sentence does not provide a clear indication of symptoms, but when considered together, they reveal relevant information. Therefore, both sentences were marked as dependent examples. Furthermore, sentences that were unrelated, such as asides or emphatic statements, were also identified and flagged. These two categories of sentences–dependent and unrelated–were subsequently removed from the dataset as they were not suitable for labeling in the context of this research question. Out of the initial 3,780 sentences, only 97 sentences (2.6%) were removed based on criteria, resulting in a dataset of 3,683 sentences. Sentences labeled as displaying signs of anxiety, depression, or both were categorized as symptomatic (positive) examples while neutral sentences served as non-symptomatic (negative) examples. The resulting training dataset was 36% symptomatic and 64% non-symptomatic sentences. Once the training data was prepared, it was shuffled and split into a typical 80% training (2,946 sentences) and 20% validation (737 sentences) dataset. The test set for evaluation purposes consisted of a clinical dataset.

**Figure 1.**
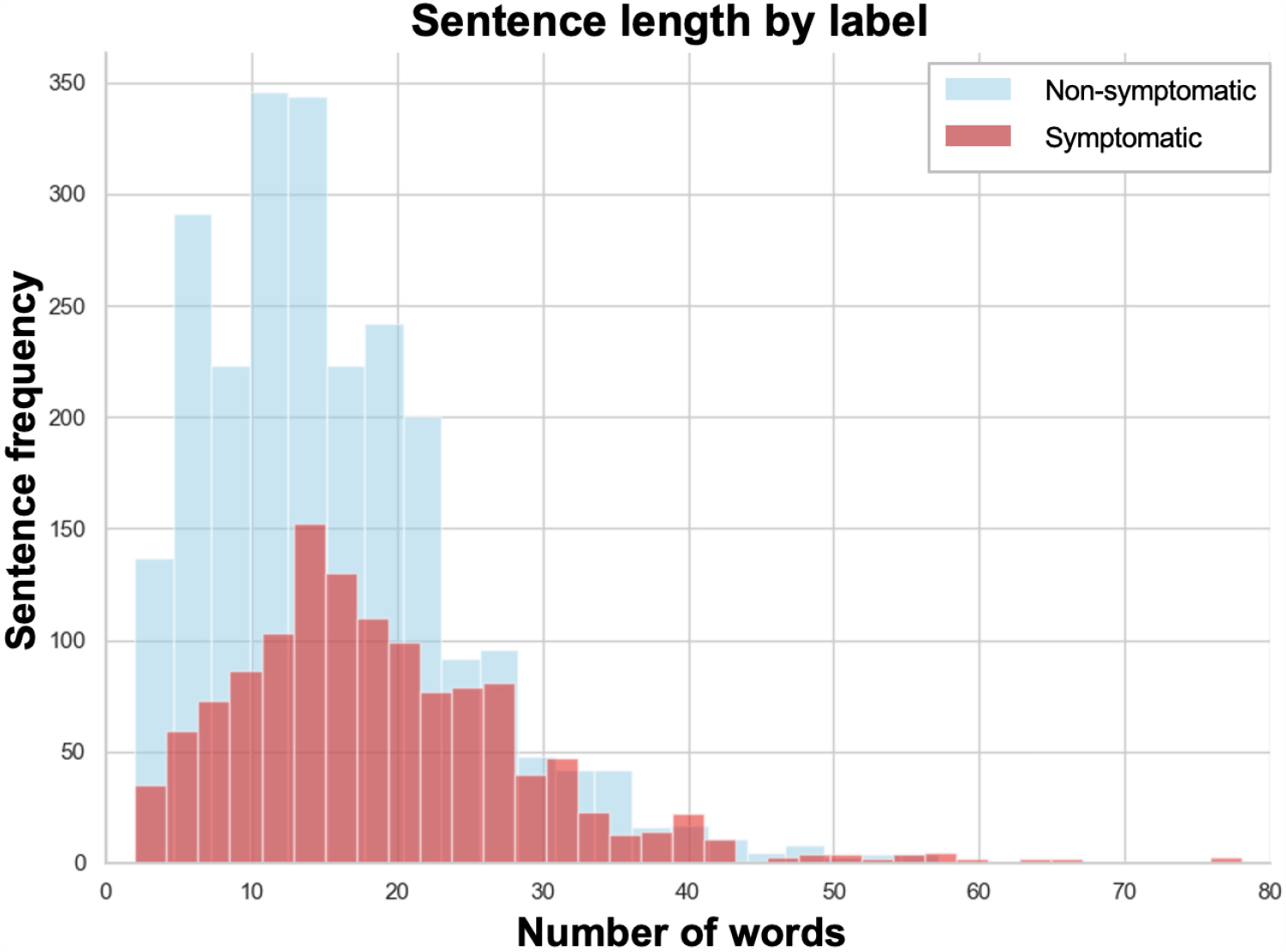
The sentence length by label type, symptomatic and non-symptomatic where symptomatic sentences were less frequent and shorter.

The training sentences used in this study had an average length of 19 words and 102 characters for symptomatic sentences, whereas non-symptomatic sentences had shorter averages of 16 words and 83 characters. In terms of data sources, each user contributed an average of 26 sentences. The highest number of sentences from a single patient in the dataset was 180 while the lowest contribution was 2 sentences.

#### 2.1.2 Testing Dataset

The clinical data utilized in this study was collected from a board-reviewed and ethically-compliant online psychotherapy clinical trial conducted at Queen’s University between 2020 and 2021. The study underwent a thorough review process by the Queen’s University Health Sciences and Affiliated Teaching Hospitals Research Ethics Board to ensure adherence to ethical standards (File #: 6020045). As part of their participation, patients provided written informed consent for the utilization of their anonymized data in academic research and publications. There were 55 subjects that participated in the trial. During the trial, participants diagnosed with major depressive disorder (MDD) received 12 sessions of therapist-supported electronic cognitive behavioral therapy (e-CBT) in asynchronous format. This asynchronous therapy involved engaging with weekly interactive online modules, which were delivered through a secure cloud-based online platform. During the initial week of the trial, participants were invited to share their personal narratives detailing their experiences with mental health challenges.

Participant narratives were segmented into 930 total sentences. Following the same inclusion criteria as the training dataset, 31 sentences (3.3%) were excluded, leaving 899 sentences for testing the algorithm performance. These sentences were similarly labeled as neutral or containing signs of anxiety and/or depression by two expert clinicians (Expert J and Expert M), who were different from the clinician involved in labeling the training dataset. Among the 899 sentences, Expert J considered 28% as symptomatic and 72% as non-symptomatic, while Expert M categorized 41.5% as symptomatic and 58.5% as non-symptomatic. Notably, this resulted in an inter-rater overlap (i.e. proportion of sentences having similar labels from both Experts J and M) of 76%, indicating a significant level of agreement between the two experts regarding sentence labeling. To assess label consistency across datasets, Expert J was also tasked with labelling the training dataset. The inter-rater overlap between Expert A and Expert J for the training dataset was 80%. These findings highlight the subjective nature of diagnostics in this field within this field.

**Figure 2.**
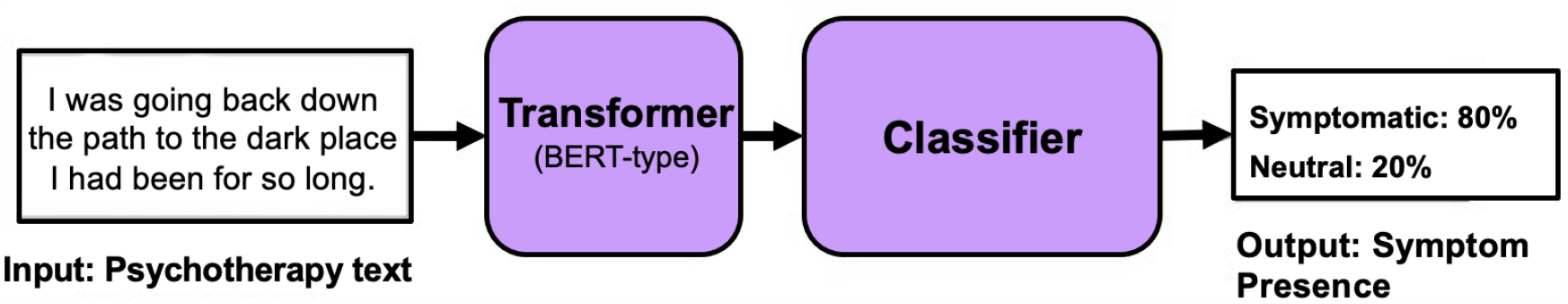
Natural language processing mental health task process figure where a narrative sentence is input to a transformer model with a classifier to predict status as either symptomatic of non-symptomatic.

### 2.2 Model Design

Due to the vast array of tasks in NLP, it is crucial to clearly define the specific task of interest prior to any modeling work. In this study, the task is a classification task, where the objective is to classify the text input based on a predefined label. Specifically, it is a binary sentence classification task, where the aim is to categorize a sentence input as one of two labels, symptomatic or non-symptomatic. Two particularly relevant subtasks in text classification are emotion recognition and sentiment analysis. Emotion recognition aims to assign a specific emotion (e.g. happy, sad, angry) to the input sentences. On the other hand, sentiment analysis focuses on capturing the overall attitude expressed in an input sentence (i.e. positive, negative, neutral). Given the nature of mental health, particularly anxiety and depression, these two subtask categories exhibit strong interrelationships and relevance to the current study.

### 2.3 Model Training

The Transformer model class was selected due to its superior performance in NLP tasks related to emotion detection, surpassing previous models that lacked contextual understanding [9, 26]. A number of models were selected from the HuggingFace transformer model library, including a standard Bidirectional Encoder Representations from Transformers (BERT) model as well other BERT-based models including RoBERTa [20], DistilBERT [23], ALBERT [18], DeBERTa [13], and XLM-RoBERTa [8]. BERT was chosen as it has established itself as the de facto and widely adopted baseline for NLP experiment [22]. In addition to training the aforementioned baseline models, training was conducted on commonly employed baseline model variants, such as cased and uncased versions, as well as large-sized models, ensuring a comprehensive exploration of the model landscape.

A selection of transformer models that had undergone further fine-tuning for text classification and specific subtasks were also included in the initial model selection phase. While models from subtasks relevant to the current task (emotion recognition, sentiment analysis) were of particular interest, a wide range of subtasks were considered. These models were chosen based on their popularity within the HuggingFace community (as determined by the number of downloads) and their demonstrated performance in various tasks. A total of 44 unique models were tested using a standardized set of hyperparameters. Each model was trained for 5 epochs. The remaining training hyperparameters were set to their default values provided by HuggingFace.

The majority of the models (75%) were baseline models that had been fine-tuned to another task before our training. Among these fine-tuned models, 57% were specifically fine-tuned for a text classification task, 16% for token classification, and 2% for fill mask tasks. The token classification models were exclusively tuned for name entity recognition (NER) subtasks while the fill mask task was for a biological subtask. The text classification models were primarily trained on subtasks of sentiment analysis (36%) and emotion recognition (32%) subtasks.

**Figure 3.**
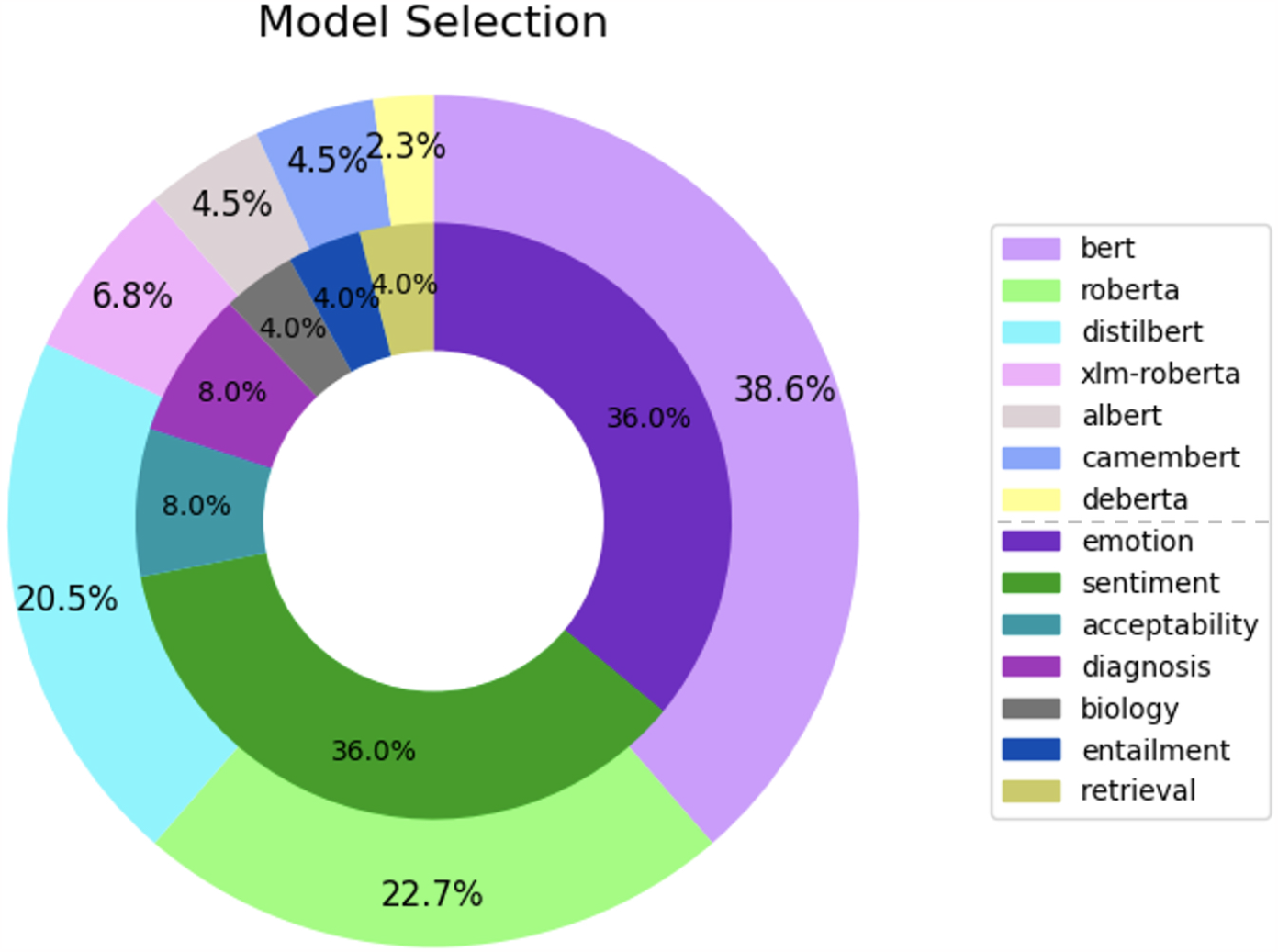
Task and subtask distributions of models used in model selection. BERT models pretrained on an emotion recognition task prior to current study training.

Prior to model training, the training examples underwent tokenization using HuggingFace’s tokenizer class, which employs base model-specific (i.e. BERT, DistilBERT, RoBERTa) tokenization techniques. It is worth noting that the training data exhibits an imbalance, with only 36% of sentences meeting the criteria for being symptomatic. To address this imbalance, a weighted cross-entropy loss function was employed during model training, where the weights were determined based on the distribution of the two classes. Model accuracy was used as the sole criteria for model selection due to the sensitive nature of the given task, which necessitates maximizing correct predictions even if it comes at the expense of computational efficiency, such as model size or latency. Specifically, the F1 score was used as the model accuracy metric, as it maintains a balance between precision and recall. For the given task, it is crucial to try to accurately predict as many symptomatic cases as possible (recall) while also maintaining a high level of confidence in the positive predictions (precision). Therefore, both metrics were considered essential in evaluating the model performance.

Data augmentation serves as a valuable approach to enhance the training dataset by introducing additional examples through slight modifications of existing ones. While in image datasets this can be achieved through simple techniques such as scaling, rotation, or color manipulation, text datasets require methods that preserve sentence meaning. Simple text augmentations like random word swap or insertion are not sufficient for complex tasks such as emotion detection, where sentence meaning plays a pivotal role. Therefore, alternative methods that better preserve sentence meaning were considered and compared, including the use of back translation. These methods offer more effective ways to generate synthetic training examples that contribute meaningfully to the training process.

Back-translation, the technique employed in this study, involves translating the original sentence to another language then back to English. It was implemented here with the NLPAug library for textual augmentation, utilizing HuggingFace transformer-based translation models. This approach is expected to maintain the sentiment of the sentence more accurately. The number and type of language intermediates used in the translation process were treated as additional tunable hyperparameters. Each available language intermediate on HuggingFace (as of 2022) was individually tested, effectively doubling the size of the training dataset with the inclusion of the back-translated synthetic counterparts for each sentence. Moreover, various ratios of back-translated synthetic sentences to the original sentence were explored, along with different combinations of language intermediates. A comprehensive investigation was conducted across 20 individual languages, followed by experimentation with the top 11 performing models in different combinations on the complete dataset. The number of languages used was incrementally increased until a drop in performance was observed. For details on this analysis, consult Appendix 1.B.

**Figure 4.**
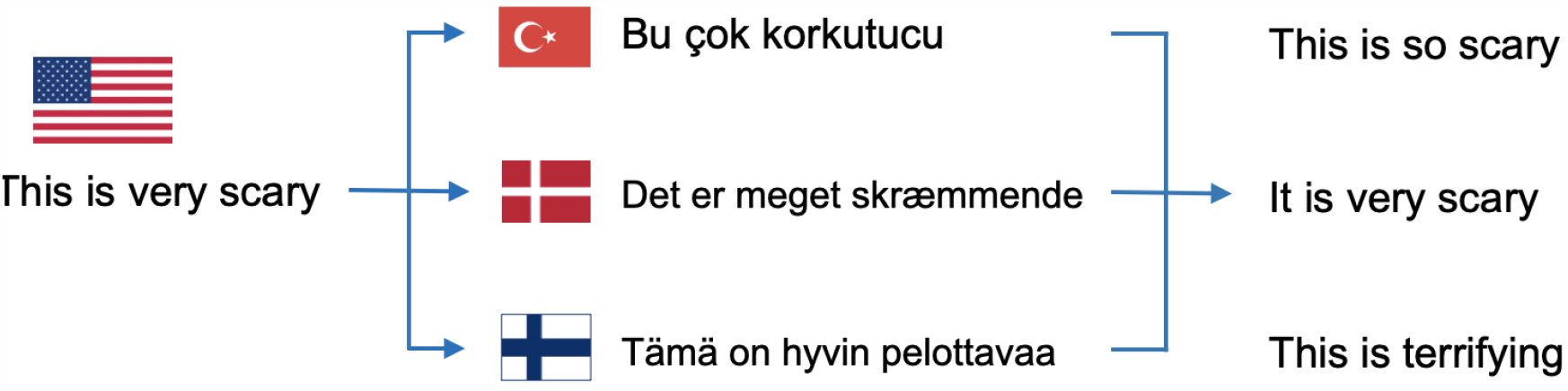
Back-translation method of data augmentation. An English sentence is translated to another language – here Turkish, Danish, and Finnish – then translated back to English with slight variations.

**Table 1:** Performance of trained models across datasets.

|  | Hyperparameter-tuned | Augmented data | Fine-tuned | fastText* |
| --- | --- | --- | --- | --- |
| Public dataset | F1: 79.3<br>bA: 84.0 | F1: 79.1<br>bA: 83.6 | F1: 77.2<br>bA: 82.3 | F1: 68.1<br>bA: 75.2 |
| Clinical Dataset (Expert J Labels) | F1: 73.0<br>bA: 72.0 |  |  |  |
| Clinical Dataset (Expert M Labels) | F1: 75.0<br>bA: 75.0 |  |  |  |

Hyperparameter tuning was conducted using the Tree Parzen Estimation (TPE) method with HyperOpt, employing an Asynchronous Successive Halving (ASHA) scheduling algorithm implemented with Ray Tune [6, 19]. The selected hyperparameters and their respective ranges for tuning were chosen based on their ability to significantly enhance model performance in prior works [24]. The hyperparameters chosen included the number of training epochs, the random seed, the number of training examples per batch, and the learning rate. The final hyperparameters used can be found in Table 5.

The fine-tuned model trained on non-clinical data was then evaluated on the clinical dataset to assess its diagnostic capabilities. The model’s performance was calculated and compared against the labels of Experts J and M, and the results are presented in Table 1.

\**All other non-DL models performed worse than fastText. For more details, please refer to the appendix*.

## 3 Results

Performance evaluation was conducted across 44 different models by comparing their F1score (harmonic mean of precision and recall) and balanced accuracy (bA) on the training dataset. The top 11 models, based on their performance, were fine-tuned using the augmented training dataset. A detailed comparison of these models can be found in Table 5.A.1 in the appendix.

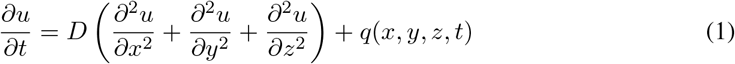

**Figure 5.**
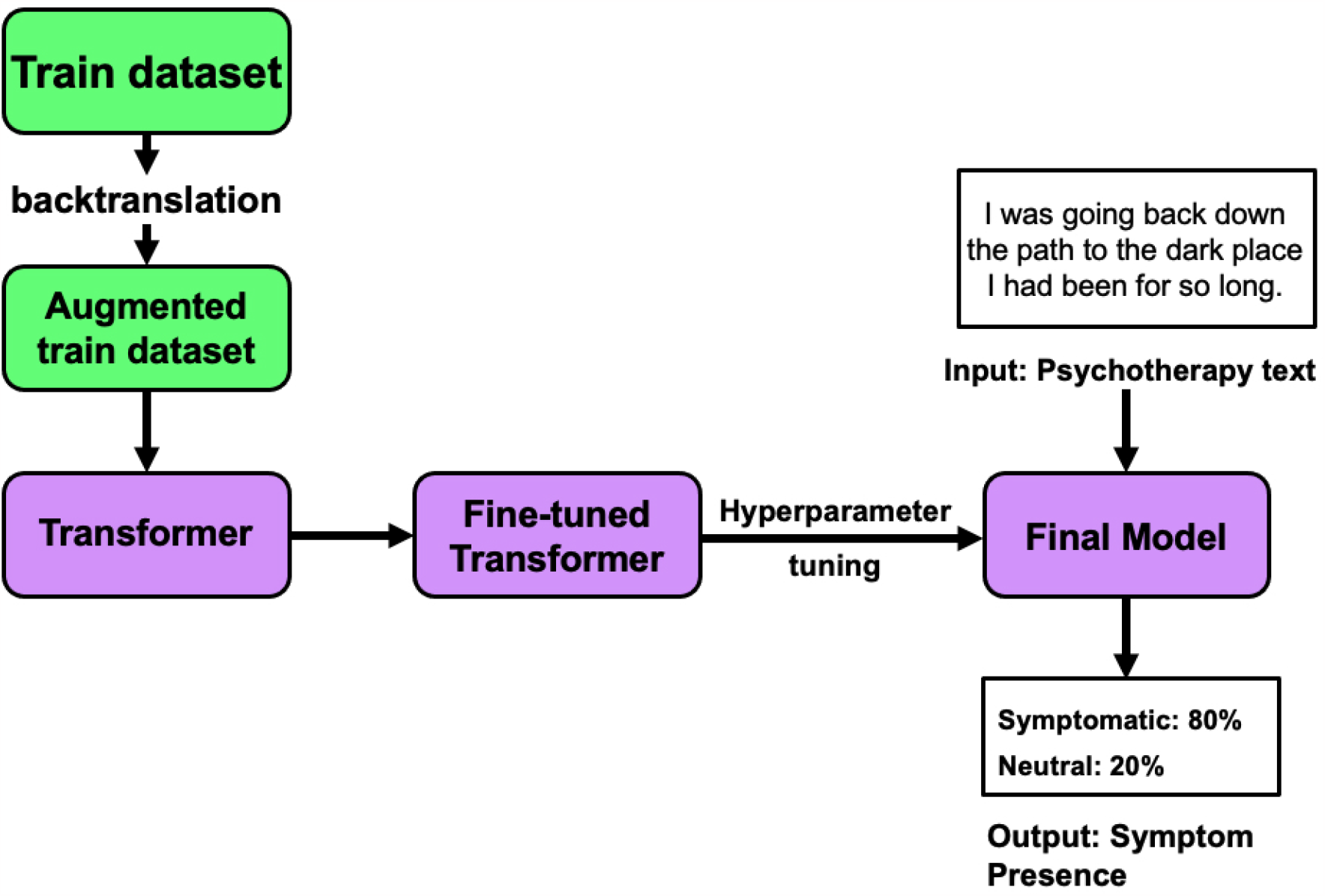
Schematic of overall training process. Train dataset is augmented then used to fine-tune a transformer model. The model is optimized through hyperparameter tuning before the final model is attained.

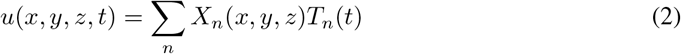

The model that achieved the highest performance in our evaluation was a fine-tuned RoBERTa model that was previously trained on the TweetEval [5] dataset prior to this dataset.^1^ TweetEval is a dataset specifically designed for multi-class emotion recognition in Tweets, consisting of over 5000 Tweets categorized into four emotions: anger, joy, sadness, optimism. By leveraging this dataset, the RoBERTa model demonstrated superior performance, outperforming word-based, context-free algorithms like fastText by approximately 10%. For a detailed analysis of each model’s performance, please consult Appendix 1.A.

Subsequently, we assessed the performance of the best-performing model, trained on the public dataset, using the clinical dataset that was labeled by two additional experts. The model achieves an accuracy of approximately 74%. It is worth noting that even human experts do not exhibit complete consistency. The inter-rater overlap between Experts J and M was found to be 76%, which is comparable to the accuracy of the model.

The results table displays results for the baseline model, which refers to the Emotion RoBERTa model. The Augmented column refers to the Emotion RoBERTa model applied to the augmented training dataset. Finally, the Tuned performance is after hyperparameter tuning.

Both the F1 score (F1) and balanced accuracy (bA) are crucial metrics included in our evaluation. These metrics are specifically employed for classification tasks involving imbalance datasets, such as this. It is important to highlight that the F1 score maintains a balance between precision and recall, whereas the balanced accuracy assess specificity and recall. The balanced accuracy metric directly considers true negatives, making it particularly useful when both true positives and true negatives are equally significant. On the other hand, the F1 score prioritizes the positive cases, emphasizing the accuracy of identifying positive instances. By utilizing both metrics, we ensure a comprehensive assessment of the model’s performance in handling imbalanced classification scenarios.

## 4 Discussion

In this project, we conducted fine-tuning on a variety of transformer models to identify symptomatic sentences in a client’s mental health narrative, specifically those related to depression and anxiety. We aimed to compare the performance of these models and address the limited availability of labeled data by employing augmentation techniques to expand our dataset.

Our findings demonstrate that our most effective model achieved an impressive accuracy of approximately 80% (F1: 79.3%, bA 84.0%) when distinguishing between symptomatic and non-symptomatic sentences, which is comparable to the performance of human experts. Despite being trained on a public dataset, this model showcased a similar level of accuracy when classifying sentences from a clinical dataset collected from 55 patients participating in a separate clinical trial (average F1=74%, bA=73.5%). Notably, our model’s performance is in line with the level of agreement between different expert raters, indicating its reliability.

Through the fine-tuning of various transformer models, augmentation techniques, and comparative analyses, our project has successfully developed a model capable of accurately classifying symptomatic sentences. Its accuracy on both public and clinical datasets, combined with its performance comparable to interrater agreement, highlights its effectiveness and potential for practical applications.

Our study aimed to identify the most effective model for the given task, and our analysis revealed that the RoBERTa model, fine-tuned on the TweetEval benchmark, outperformed the other models examined. RoBERTa is an enhanced version of the original BERT transformer model, benefiting from robust optimization during pretraining, which ultimately resulted in improved model performance [9, 20]. Unlike BERT, RoBERTa underwent pretraining using an expanded dataset, comprising of five English-language corpora that totaled over 160 GB of uncompressed text. These corpora include BOOKCORPUS [29], WIKIPEDIA, CC-NEWS [21], OPENWEBTEXT [11], STORIES [25].

The model was further fine-tuned on the TweetEval benchmark prior to our task. Specifically, it utilized the Emotion Recognition dataset, which contains of over 5000 text statements sourced from Twitter. Each statement is associated with one of four emotions: anger, joy, sadness, and optimism. The exceptional performance of the RoBERTa model highlights the significance of emotion classification as a valuable precursor for mental health diagnostics. It is worth noting that the BERT base model, XLM-RoBERTa pretrained on a sentiment analysis task, and DistilBERT base models closely trailed in terms of performance.

By highlighting the superior performance of the RoBERTa model fine-tuned on Emotion TweetEval, our study underscores its efficacy as the optimal choice for the given task. This model’s success indicated the potential utility of emotion classification in facilitating health diagnostics.

One effective strategy for enhancing model training involves employing diverse data augmentation techniques to expand the training dataset. In our study, we explored the back-translation technique and observed a notable improvement in model performance as a result. Interestingly, we discovered that using intermediate languages from the Indo-European, Turkic, or Uralic language families yielded superior results compared to other language families, such as Sino-Tibetan, Japonic, Austronesian, or Afro-Asiatic. This can be attributed to the fact that languages belonging to the same language family as English tend to capture sentence structure and meaning more effectively due to their greater similarity.

To maximize the benefits of back-translation, we identified the combination of Turkish and Danish as particularly effective, allowing us to triple the size of the training dataset by generating two additional augmented sentences for each origin training sentence. This language combination produced the highest model performance, achieving an approximate 2% increase in the F1 score. For a detailed analysis of the language combination, corresponding performance, and the impact of increasing the ratio of synthetic sentences to original sentences, please refer to the appendix A2, where a table is provided.

By strategically implementing back-translation and specifically leveraging the Turkish and Danish languages, we successfully amplified the training dataset and enhanced the model’s performance. The observed improvements validate the effectiveness of this approach for training models for the given task.

During the hyperparameter tuning process, we focused on optimizing several key hyperparameters, including the number of training epochs, random seed, number of training examples per batch, and the learning rate. While these hyperparameters were carefully tuned, it is worth noting that there may still be room for further optimization. After thorough experimentation, we observed only a modest improvement in the overall model performance, with the F1 scores increasing by a mere 0.2%. For a comprehensive list of the final set of hyperparameters employed in this study, please refer to the appendix A3. Moving forwarded, further exploration and optimization of hyperparameters may garner additional model success. It is an avenue that warrants future investigation and can potentially yield more substantial improvements.

The findings from this study highlight the promising potential of training a transformer model for a nuanced and intricate clinical task, specifically the detection of symptomatic language use, even when faced with limited labeled data. Furthermore, the transferability of the model’s knowledge to diverse datasets collected in distinct clinical settings is a crucial outcomes. Ultimately, these transformer models have the capacity to revolutionize the field by enabling scalable and objective mental status evaluations based on patients’ language usage.

As we move forward, it becomes imperative to envision future clinical trials that leverage these objective measurements to predict essential clinical outcomes. These outcomes could expand to include factors such as patient engagement, symptom reduction, or even relapse prediction. By incorporating these objective measurements into the design of future trials, we can potentially enhance our understanding of the complex interplay between language use and clinical outcomes.

By utilizing the power of transformer models and their ability to accurately analyze language patterns, we pave the way for more precise and comprehensive evaluations of mental health. This has the potential to significantly impact clinical practice and improve patient care. The next critical step is to strategically integrate these models into clinical trials, enabling the generation of invaluable insights that can inform treatment decisions and interventions, ultimately enhancing patient outcomes and well-being.

## Data Availability

All data produced in the present study are available upon reasonable request to the authors.

## Appendix

### A.1 Model Selection

Table 2 displays the top 5 performing transformer models using F1 score as the sorting metric. These scores are without data augmentation, hyperparameter tuning, or any other optimizations. Balanced accuracy is seen to monotonically increase as well as F1 score. Interestingly, if the models are sorted by balanced accuracy performance, the top four models remain the same, and the fifth best model swaps with the sixth best model (F1: 76.50% BA: 81.72%, model type: RoBERTa). Three of the top five models are a RoBERTa model or model variant. Two are baseline models while three are models fine-tuned on tweets for another related emotion or sentiment task.

**Table 2:** Performance of trained models across datasets.

| Model Name (*source*) | Model Type | F1 Score | Balanced Acc |
| --- | --- | --- | --- |
| TweetEmotionEval (*elonzo*) | RoBERTa | 77.26% | 82.37% |
| BERT base uncased | BERT | 77.04% | 82.00% |
| TwitterSentiment (*cardiffnlp*) | XLM-RoBERTa | 76.92% | 81.95% |
| DistilBERT base uncased | DistilBERT | 76.81% | 81.81% |
| TwitterSentiment (*cardiffnlp*) | RoBERTa | 76.52% | 81.60% |

Nine of the forty-eight models performed no better than chance, with an F1 and BA of 0.5 and were thus removed from the following analyses as outliers.

**Figure 6.**
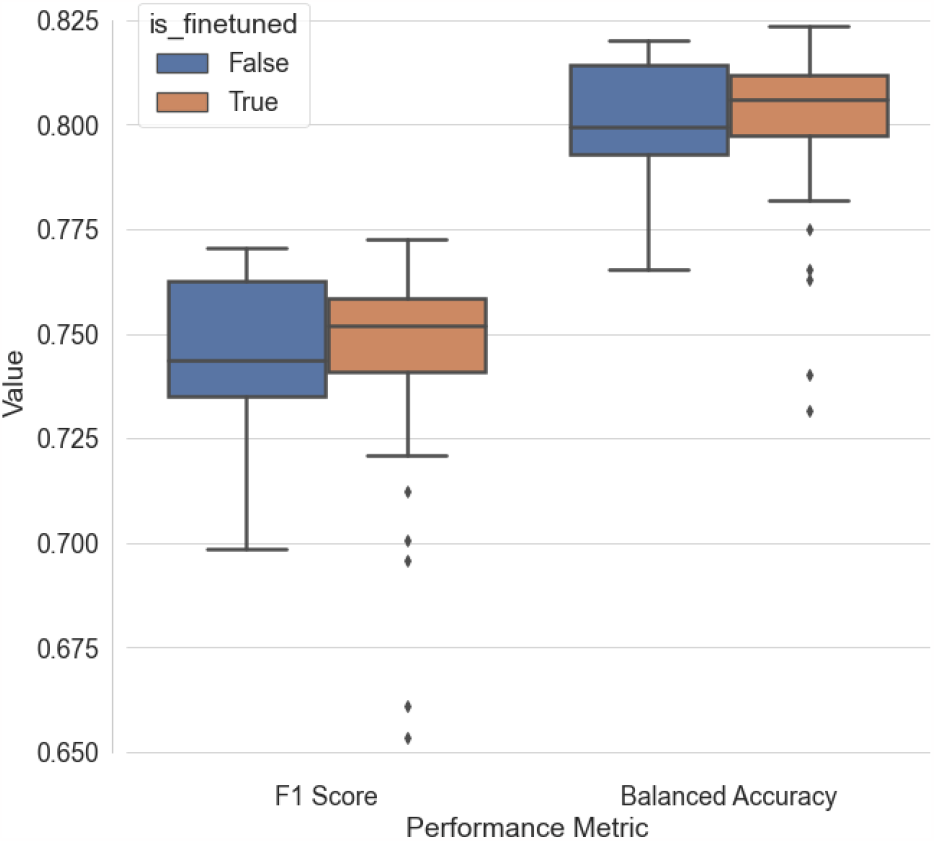
Performance of all transformer models trained during model selection. Models previously fine-tuned on related subtasks (*orange*) tended to perform slightly better on average than base models (*blue*).

**Table 3:**
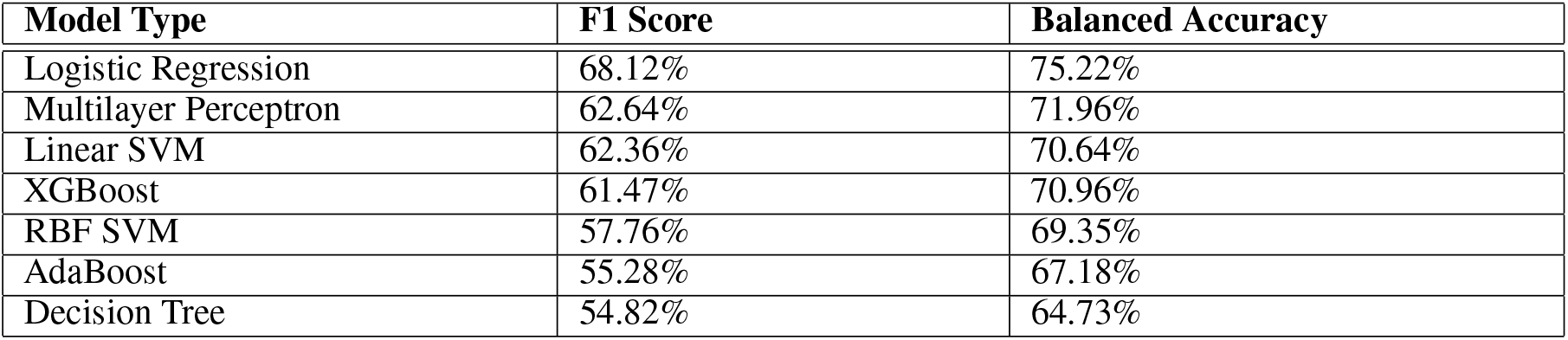
Performance of Classical Machine Learning Models.

### A.2 Back-translation Data Augmentation Model Performance

**Table 4:** Performance of top 5 language combinations.

| Languages | F1 Score | Balanced Acc |
| --- | --- | --- |
| Turkish + Danish | 79.10% | 83.64% |
| Bengali + Turkish + Finnish + Spanish | 78.55% | 83.38% |
| Bengali + Uralic + Finnish + Portuguese | 78.24% | 83.11% |
| Bengali + Turkish + Dutch + Estonian | 78.18% | 83.09% |
| Spanish | 78.15% | 82.97% |

**Table 5:**
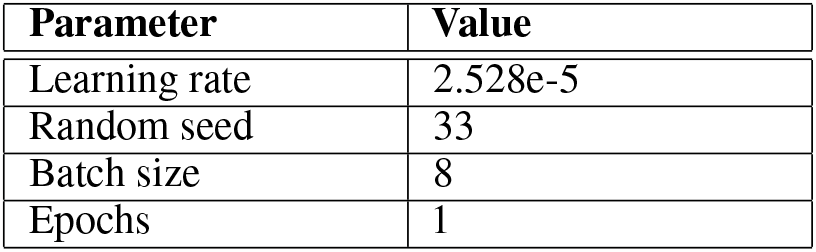
Final hyperparameter values after tuning.

**Figure 7.**
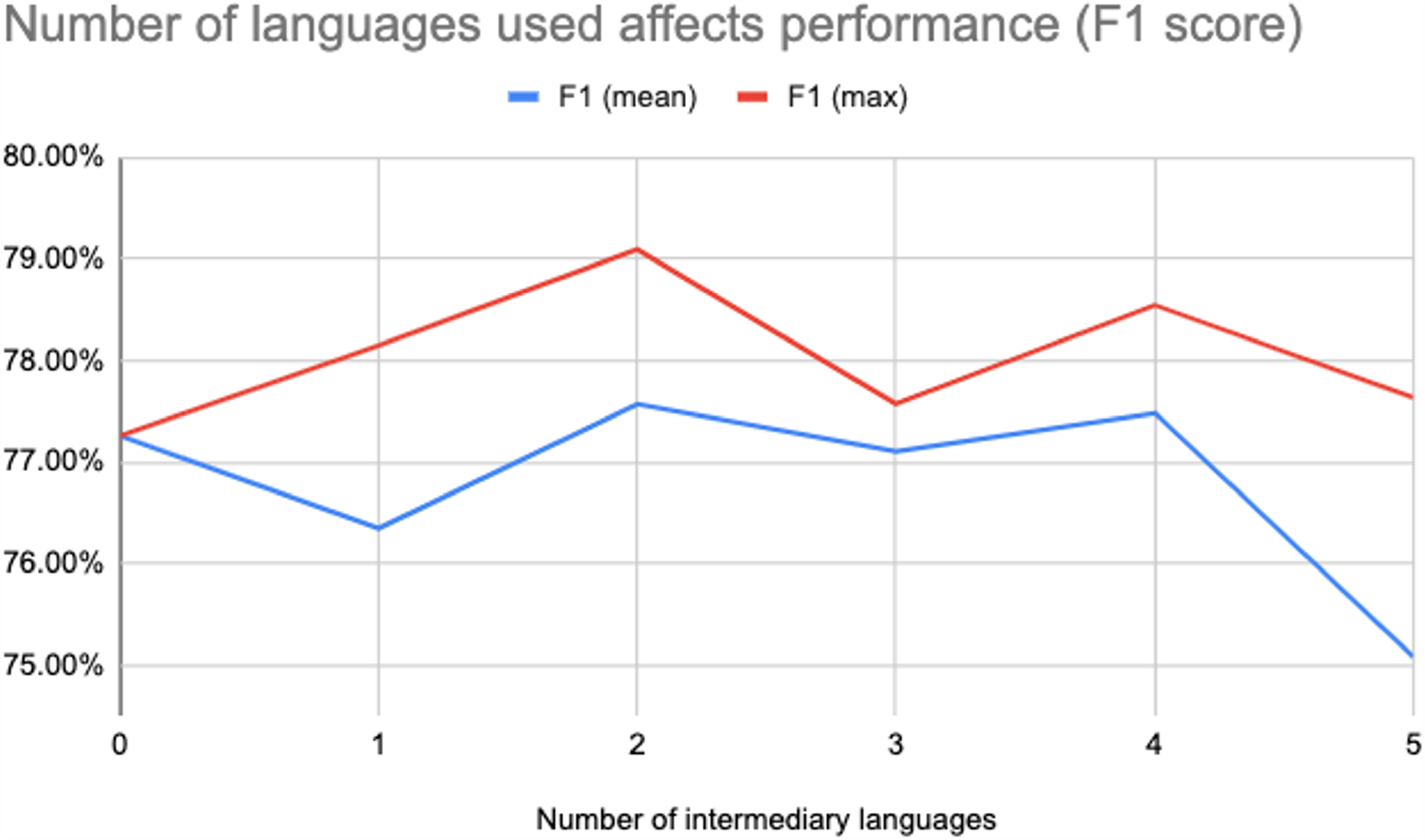
Effect of number of intermediary languages on model performance in back-translation data augmentation. Models performed best with two intermediary languages and worse with five or more languages.

### A.3 Final Hyperparameters

## Acknowledgments and Disclosure of Funding

Use unnumbered first level headings for the acknowledgments. All acknowledgments go at the end of the paper before the list of references. Moreover, you are required to declare funding (financial activities supporting the submitted work) and competing interests (related financial activities outside the submitted work).

## Footnotes

1 The model has been made available on HuggingFace as margotwagner/roberta-psychotherapy-eval

## Notes

### Competing Interest Statement

The authors have declared no competing interest.

### Funding Statement

This study was funded by

### Author Declarations

Research Ethics Board of Queens University Health Sciences and Affiliated Teaching Hospitals gave ethical approval for this work

